# Hospital admissions for respiratory tract infections in children aged 0-5 years for 2017/2023

**DOI:** 10.1101/2021.11.22.21266564

**Authors:** Fredrik Methi, Ketil Størdal, Kjetil Telle, Vilde Bergstad Larsen, Karin Magnusson

## Abstract

**Aim:** To compare hospital admissions across common respiratory tract infections (RTI) in 2017-21, and project possible hospital admissions for the RTIs among children aged 0-12 months and 1-5 years in 2022 and 2023.

**Methods:** In 644 885 children aged 0-12 months and 1-5 years, we plotted the observed monthly number of RTI admissions (upper- and lower RTI, influenza, respiratory syncytial virus (RSV), and COVID-19) from January 1^st^, 2017 until October 31^st^, 2021. We also plotted the number of RTI admissions with a need for respiratory support. We used the observed data to project four different scenarios of RTI admissions for the rest of 2021 until 2023, with different impacts on hospital wards: 1) “Business as usual”, 2) “Continuous lockdown”, 3) “Children’s immunity debt”, and 4) “Maternal and child immunity debt”.

**Results:** By October 31^st^, 2021, the number of simultaneous RTI admissions had exceeded the numbers usually observed at the typical season peak in January, i.e. ∼900. Based on our observed data and assuming that children and their mothers (who transfer antibodies to the very youngest) have not been exposed to RTI over the last one and a half years, our scenarios suggest that hospitals should be prepared to handle *two to three times* as many RTI admissions, and *two to three times* as many RTI admissions requiring respiratory support among 0-5-year-olds as normal, from November 2021 to April 2022.

**Conclusion:** Scenarios with immunity debt suggest that pediatric hospital wards and policy makers should plan for extended capacity.

## Introduction

The lockdown restrictions that were implemented to control the transmission of the SARS-CoV-2 have likely lowered the incidence of COVID-19 and hospital admissions with COVID-19 in Norway. The restrictions may also have controlled the transmission of other respiratory tract infections (RTI) that are common in children, i.e., transmission of respiratory syncytial virus (RSV) or influenza viruses [1-5]. Consequently, children may have been less exposed to viruses causing RTI during the COVID-19 pandemic. Lack of exposure to mothers during pregnancy can add to the youngest children’s vulnerability, as maternal antibodies may be lacking. In a normal season, 60-70% of all children admitted with RSV infection are younger than 12 months, and 80% are younger than two years [6-7]. The lack of exposure during the pandemic lockdown has an unknown impact on their young and developing immune system. We hypothesize that the pandemic and its accompanying restrictions have resulted in an immunity debt among the youngest children and their mothers, which will lead to a sharp rise in hospitalizations for children aged 0-5 (with or without need for respiratory support) for all RTI infections during the RTI season 2021/22. The aim of the study is to provide knowledge which can help pediatric hospital wards to better foresee capacity needs in the upcoming RTI season.

## Methods

### Design & data sources

Beredt C19 is a nation-wide emergency preparedness register aiming to provide rapid knowledge about the pandemic, including impacts of measures to limit the spread of the virus on health and utilization of health care services [8]. Beredt C19 compiles daily individual-level data from several registers, most important for the current study: the Norwegian Patient Register (NPR) (all electronic patient records from all hospitals in Norway) and the National Population Register (age, sex). The establishment of an emergency preparedness register forms part of the legally mandated responsibilities of The Norwegian Institute of Public Health (NIPH) during pandemics. Institutional board review was conducted, and The Ethics Committee of South-East Norway confirmed (June 4^th^, 2020, #153204) that external ethical board review was not required.

### Population

Our population included all children aged 0-5 years who were Norwegian residents during the period from January 1^st^, 2017 to September 30^th^, 2021. Children who were born (stillbirths not included) or who immigrated were included in both the numerator and denominator from the first full month following the date of birth or immigration (and similarly excluded in the month of dying or emigrating, which was extremely rare).

### Outcome: RTI and COVID-19

We studied all inpatient hospital contacts independent of length of stay and urgency. Hospital contacts that occurred with less than 48 hours in between were coded as the same admission. In cases where multiple diagnostic codes were registered, we selected the code with the highest specificity (i.e., with known pathogen if available). Outpatient contacts occurring less than 48 hours before or after admission were included as the same admission. We studied the prevalence of hospital admissions in 66 International Classification of Diseases (ICD-10) codes related to RTI [9]. Moreover, we divided these into five mutually exclusive categories (S-Table 1): Upper RTI, lower RTI, influenza-coded RTI, RSV-coded RTI and COVID-19. Further, to shed light on the severity grade of the different diseases leading to hospitalization, we studied the categories if combined with respiratory support (S-Table 1).

### Statistical analyses

First, we plotted the observed monthly number of all RTI admissions from January 1^st^, 2017 until October 31^st^, 2021. Second, we estimated the number admitted per month in a simple model with a linear monthly trend and binary variables for each calendar month from January 1^st^, 2017 until December 31^st^, 2019 and used this model to predict the number admitted per month from August 2021 until June 2023. We started the projections from August 2021 to capture all the immunity debt. The model was mended using the following assumptions to illustrate four possible scenarios with different impacts on the health services:

#### “Business as usual”

The number of children admitted for the different RTIs *will not be affected* by the pandemic and will from November 2021 and onwards follow the same seasonal pattern as observed in 2017-19 by calendar month. Hence, in this scenario, we assumed that the drop in the number of RTI admissions during the pandemic was only temporary.

#### “Continuous lockdown”

The number of children admitted for the different RTIs *will be affected in the same way* as during the pandemic lockdown measures, and will from November 2021 and onwards follow the same seasonal pattern as observed between July 2020 and June 2021. Thus, we here assumed that the impact of lockdown measures is permanent.

#### “Children’s immunity debt”

Children have not been exposed to RTI viruses over the last two years, presumably resulting in excess susceptibility due to lack of immunity [10]. Thus, the number of children admitted for the different RTIs *will be temporarily affected*, assuming that every hospitalization for any of the RTIs that did not occur throughout the lockdown period (i.e., 2020-21) will occur in the upcoming season (2021-22) before gradually stabilizing in the same pattern as described in the “business as usual”-scenario. Thus, in addition to the hospitalizations in ‘business as usual’ from November 2021 onwards, we assumed that the hospitalizations that normally would have occurred between July 2020 and June 2021 will instead occur from November 2021 to June 2022.

#### “Maternal and child immunity debt”

Pregnant mothers have not been exposed to RTI viruses over the last one and a half years, implying that fewer antibodies were transferred from mothers to offspring through placenta and breastfeeding [11-12]. It has previously been found that children being breastfed ≤ 6 months have a 25% increased risk of being infected with RTI compared to those being breastfed ≥ 12 months [13]. This may result in an excess susceptibility among the newborn or breastfed children (here defined as children aged 0-12 months). Thus, in addition to the seasonal RTI hospitalizations observed in 2017-19 and the immunological debt from 2020-21 (i.e. in accordance with the “children’s immunity debt”-scenario), the number admitted for the different RTIs may *be temporarily severely affected*, and we assumed twice as many RTI hospitalizations in 2022 among the 0-12-month-olds. This scenario is the same as the “children’s immunity debt”-scenario, except that the number of hospitalizations among 0-12-month-olds is doubled.

We repeated the analyses under the four scenarios using the diagnostic groups in combination with respiratory support as outcome variable. In-depth explanations of the different scenarios are available in S-Methods. All analyses were run in STATA MP v16.

## Results

We included all children aged 0 to 5 years from the first month following their birth date. In total we included 644 885 unique children, and on average we studied the number of hospital admissions among 349 649 children every month, i.e. the denominator ranged from a maximum of 363 133 children in February 2017 to a minimum of 335 535 children in September 2021. The mean (SD) age for all observations (children-month) was 3.1 (1.7) years and the sample consisted of 51% boys.

### 2017-2020 trends for the different RTI admissions

Upper- and lower RTI, RSV and influenza admissions all followed very similar seasonal patterns from January 2017 to January 2020 for children aged 0-5 years, peaking around the turn of the year (Figure 1). Among the included types of RTI admissions, upper RTI was the most common cause (217 per month on average), followed by lower RTI (106 per month), RSV (89 per month) and influenza (18 per month). Altogether, a peak of approximately 1000 children per month were admitted for upper- and lower RTI, RSV and influenza in January these years, with a significantly lower number in the summer (Figure 1, Figure 2). In contrast, COVID-19 admissions had no similar seasonal variations, fluctuating between 0 and 13 admissions per month with a monthly average of five admissions between March 2020 and October 2021 (Figure 2). The number of RTI admissions with a combined need for respiratory support was lower with a peak of approximately 100-200 admissions per month in January 2017-19 (Figure 1, Figure 2). As expected, the numbers of RTI respiratory support admissions followed the same seasonal variations as the number of all RTI admissions (Figure 1, Figure 2).

**Figure 1.**
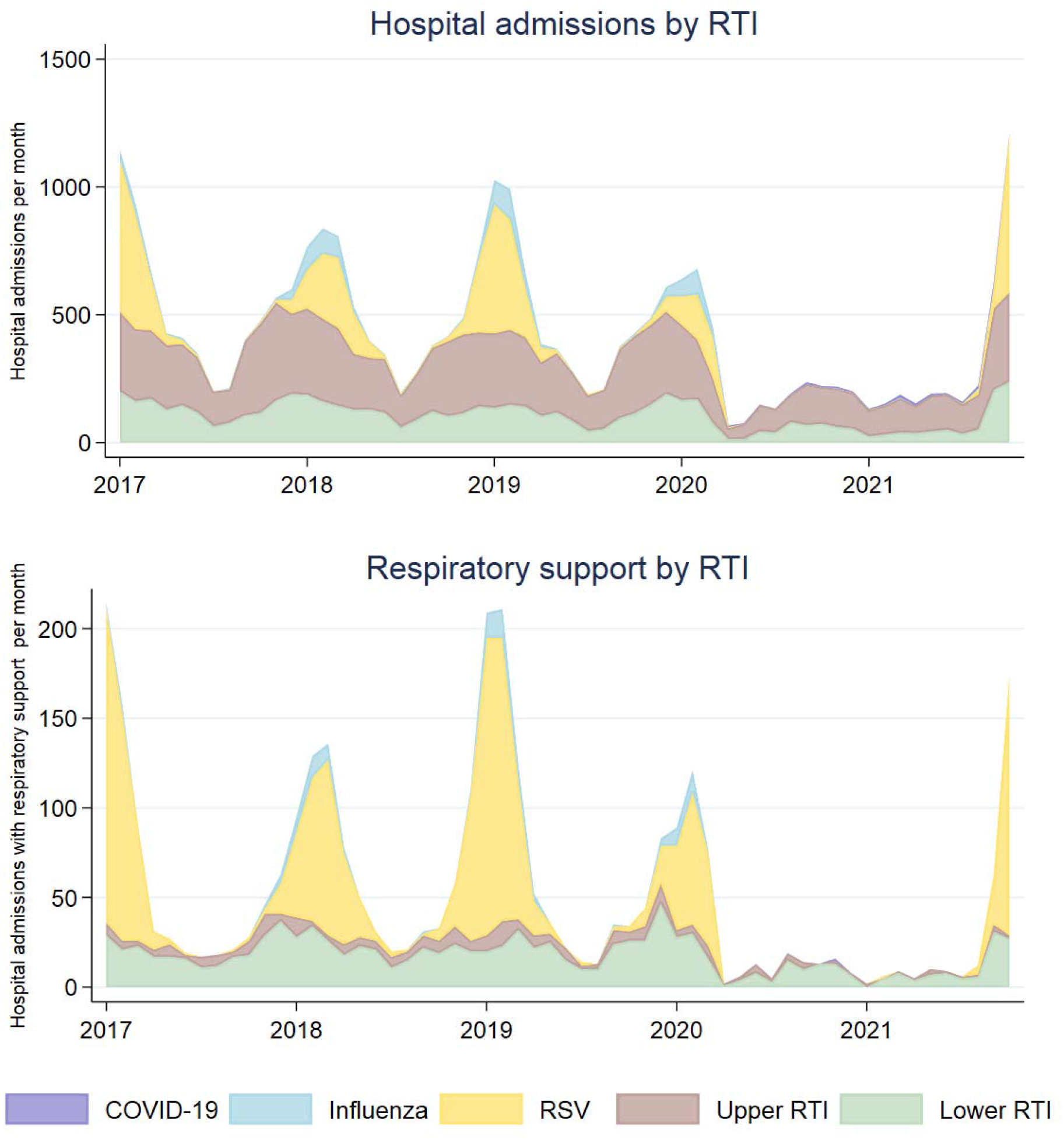
Observed number of monthly respiratory tract infections (RTI) resulting in hospital admission (upper panel) and in need for respiratory support (lower panel) for children aged 0-5 years for the five mutually exclusive groups COVID-19, influenza, respiratory syncytial virus (RSV) and upper- and lower unspecified RTI in Norway, January 1^st^, 2017-October 31^st^, 2021. The number of COVID-19 cases were <100 with a monthly peak of 15 and are barely visible in either panel.

**Figure 2.**
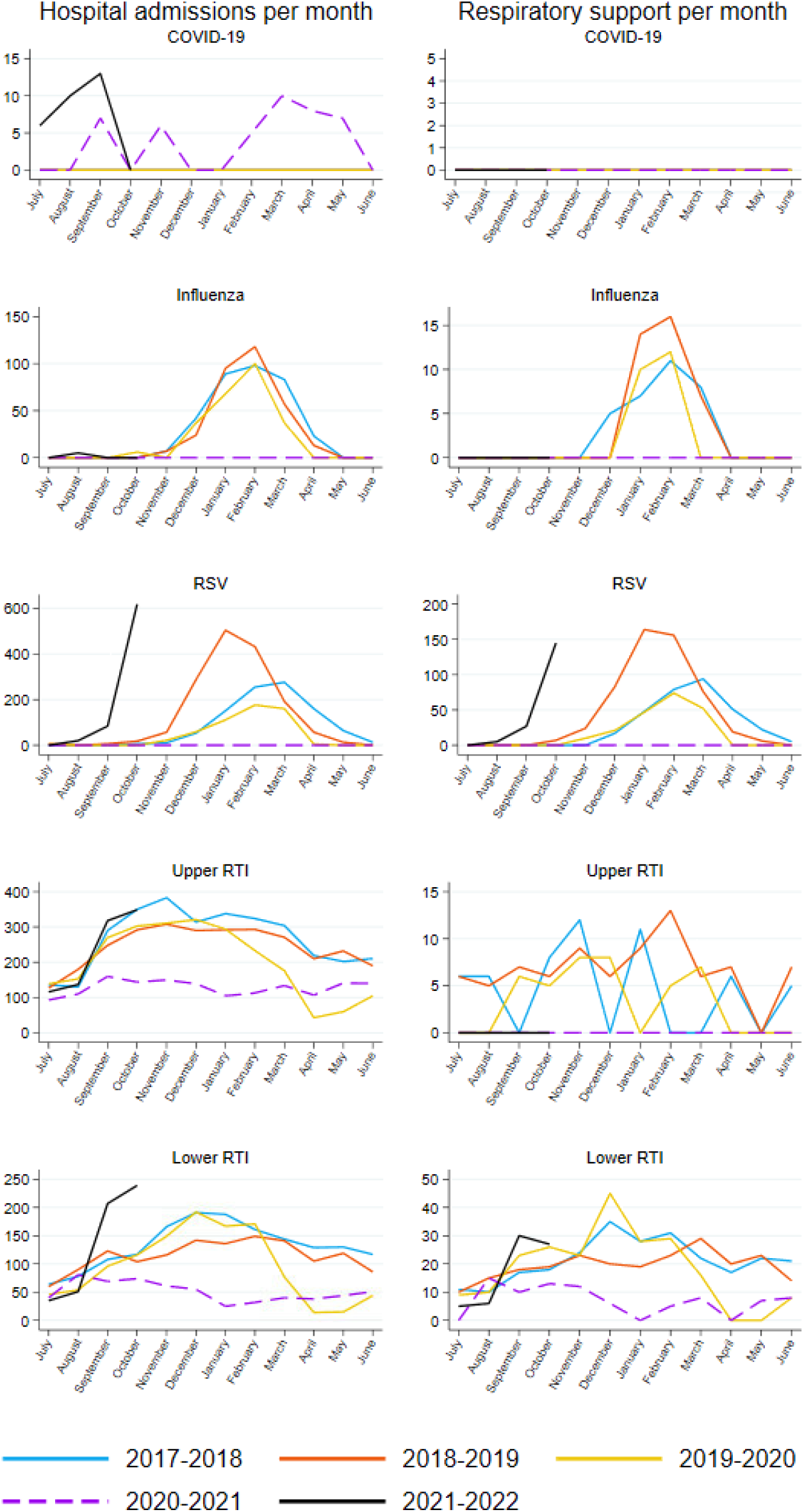
Observed number of respiratory tract infections (RTI) resulting in hospital admission for children aged 0-5 years by calendar months in Norway, January 1^st^, 2017 – October 31^st^, 2021. RSV: respiratory syncytial virus. Values < 5 are set to 0 for privacy reasons.

### Projections for 2022 and 2023

First, the scenario “Business as usual” projects a peak number of 916 RTI admissions (all five disease categories combined) in January 2022 and 196 RTI respiratory support admissions in February 2022, while the observed data shows 1211 and 174 admissions already by October 2021, respectively (Figure 3, S-Table 2). This scenario is not in line with the observed data.

**Figure 3.**
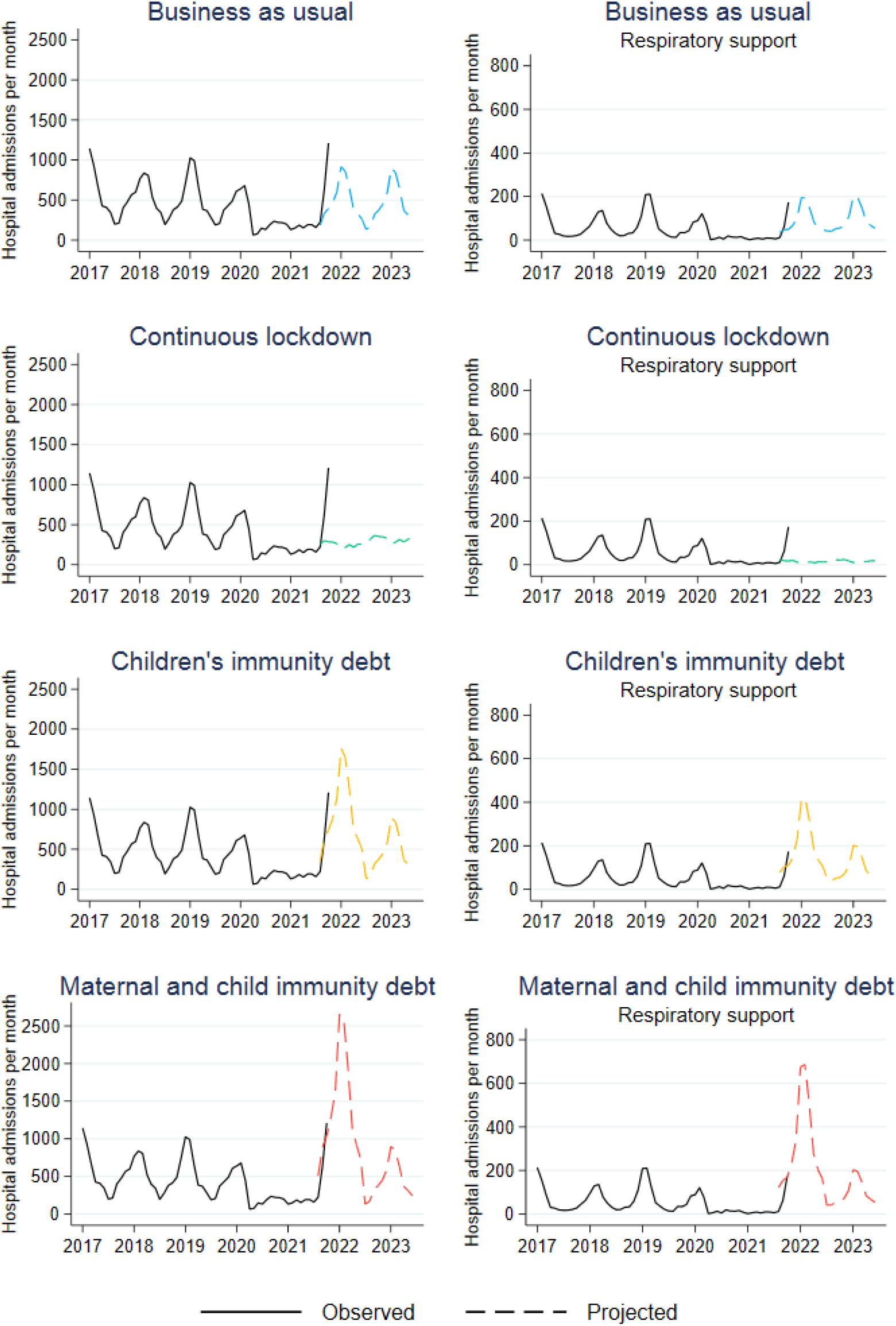
Observed (January 1^st^, 2017-October 31^st^, 2021) and projected (from August 1^st^, 2021 and onwards) number of respiratory tract infections resulting in hospital admission for children aged 0-5 years in Norway.

Second, the scenario “Continuous lockdown” projects very low numbers (peak of 366 admission and 28 in need of respiratory support) of admissions in the 2021-22 season and into the 2022-23 season. Again, this is much lower and not in line with the observed data (Figure 3, S-Table 2).

Third, the scenario “Children’s immunity debt” projects a peak of approximately 1750 RTI admissions in January 2022 and 419 RTI admissions with respiratory support in February 2022, before eventually declining back to a ‘normal’ level of 900 RTI admissions and 203 RTI admissions with respiratory support in January 2023 (Figure 3, S-Table 2). This development is largely in line with the observed data up to October 2021 and thus possibly consistent with the assumption that the disease control measures that were implemented to temporarily restrict transmission of COVID-19, have resulted in an immunological RTI debt which is paid off by 2022-23. This scenario would imply that a Norwegian hospital typically treating fifty children for RTI at a given point in peak season, should be prepared to handle the double, i.e., 100.

And finally, the scenario “Maternal and child immunity debt” projects a peak of 2,664 children admitted with RTIs in January 2022 and 686 children admitted with RTI and respiratory support in February 2022, before this eventually declines back to a ‘normal’ level of 900 RTI admissions and 203 RTI admissions with respiratory support in January 2023 (Figure 3, S-Table 2). This is also in line with the observed data up to October 2021, and thus possibly consistent with assumptions that children, especially newborn and breastfed children, lack immunity to the extent that the number of hospitalizations among children aged 0-12 months will double.

## Discussion

In this study of observed and projected RTI admissions among children aged 0-5 years, we show that the number of RTI admissions by the end of October 2021 has already reached the peak number of RTI admission as observed in RTI seasons 2017 to 2019. The scenarios “Business as usual” and “Continuous lockdown” are illustrative, however they project admissions far below the observed data up to the end of October 2021. The scenarios “Children’s immunity debt” or “Maternal and child immunity debt” are in line with the observed data.

Provided that children and their mothers (infants have maternal antibodies that are transferred during late pregnancy) have not been exposed to the usual burden of RTI over the last one and a half years [10-14], our projections suggest that hospitals should be prepared to handle *two to three times* as many RTI admissions among 0-5-year-olds as in normal season peaks. Similarly, hospitals should be prepared to handle two to three times as many RTI admissions requiring respiratory support therapy during the upcoming season.

Our models are based on observed seasonal variations and strong assumptions about the development of RTI admissions among the youngest children. As such, the very simple and transparent models may function as tools for pediatric hospital wards in planning for the upcoming peak season. For example, hospital wards might need to ensure not only a sufficient number of hospital beds, but also that a sufficient number of respiratory support staff and medical equipment (CPAP, high flow oxygen therapy and ventilators) suited for children can be summoned at peak season. In that regard, the present study has some important and very urgent clinical implications as well as important health policy implications like prioritization and funding. However, the transmissibility of respiratory pathogens is not easy to foresee as they may depend on a range of factors not accounted for here (for example, the presence of different influenza viruses circulating in different proportions. Also, we made only one assumption for all RTIs as to how long mothers transfer antigens to their newborn offspring. In reality, this may vary with the different RTIs and the final scenario “Maternal and child immunity debt” may therefore be an overestimate).

Major strengths of our study are its simplicity, transparent assumptions, and immediate impact as well as the inclusion of data from all children who are hospitalized and not hospitalized in Norway from 2017 to October 2021. Important limitations are the lack of data on positive and negative tests for all RTI viruses, and the inability to make out-of-sample validation of the model since we only have data from 2017 onwards. As such, it is possible that the projected peak for 2021-22 will spread out over both 2022, 2023 and 2024 and onwards, with lower peaks and higher numbers of RTI admissions than ‘normal’, i.e. lasting for several years. It is also possible that counteractive policy measures can be taken to contain transmission of RTI viruses and thus flatten the admission curve. Similarly, an alternative scenario not included in our work, is that of a shifted peak in RTI admissions. Instead of having a peak of hospitalized children in January, we might already have observed the peak in October. Such an early peak might increase the likelihood of a second peak during the same season, which is an additional possible scenario not considered in this study. In Denmark a peak in confirmed RSV cases this year seems to have occurred already in September 2021 [4]. Sweden shows similar evidence with what may be a peak in confirmed cases for children aged 0-4 years in October 2021 [5]. A final limitation may be that our model is purely reproducing the empirical patterns of 2017-2020 and that the number of projected scenarios as well as the content in each scenario are limited. As an example, we are not accounting for possible exponential transmissions of RTI viruses and associated hospitalizations.

## Conclusion

In conclusion, and based on the observed trend from January 2017 to October 2021, hospitals and health care authorities should continue to plan for extended capacity to be able to face a potentially massive wave of RTI admissions among young children. We believe this surge in RTI infections among our most vulnerable children when COVID-19 restrictions have been eased, is an important lesson to be learnt for the handling of potential future pandemics affecting mainly the elderly.

## Data Availability

The individual-level data used in this study are not publicly available due to privacy laws. However, the individual-level data in the registries compiled in Beredt C19 are accessible to authorized researchers after ethical approval and application to helsedata.no administered by the Norwegian Directorate of eHealth. Stata do-files are available upon request.

## Acknowledgements

We would like to thank the Norwegian Directorate of Health, in particular Director for Health Registries Olav Isak Sjøflot and his department, for excellent cooperation in establishing the emergency preparedness register. We would also like to thank Gutorm Høgåsen, Ragnhild Valen and Anja Elsrud Schou Lindman for their invaluable efforts in the work on the register. The interpretation and reporting of the data are the sole responsibility of the authors, and no endorsement by the register is intended or should be inferred. We would also like to thank Pål Surén, Margrethe Greve-Isdahl and Trine Hessevik Paulsen for their invaluable feedback on the drafted manuscript. Finally, we would like to thank everyone at the Norwegian Institute of Public Health who has been part of the outbreak investigation and response team.

## Ethics approval

The establishment of an emergency preparedness register forms part of the legally mandated responsibilities of The Norwegian Institute of Public Health (NIPH) during epidemics. Institutional board review was conducted, and the Ethics Committee of South-East Norway confirmed (June 4th, 2020, #153204) that external ethical board review was not required.

## Consent

Individual consent was not required because the current study was based on routinely collected and deidentified administrative data.

## Authors’ contributions

FM takes full responsibility for the integrity of the data and the accuracy of the data analysis. FM, KT, VBL and KM had access to, and verified, the underlying data. All authors designed the study. KS, KT, and KM critically evaluated all stages of the research process. All authors contributed with conceptual design, analyses, and interpretation of the results. All authors contributed with drafting the article and critically revising it for important intellectual content.

All authors gave final approval for the version to be submitted.

