## Supplementary material for "Hospital admissions for respiratory tract infections in children aged 0-5 years for 2017/2023"

By Methi et al., 2021.

**S-Table 1:** ICD-10 codes and RTI categories. p. 2

**S-Methods:** Estimating projections. p. 3-4

**S-Table 2:** Observed vs. projected RTI-admissions + respiratory support. p. 5

**S-Table 1:** ICD-10 codes and RTI categories

| Categories | ICD-10 codes |
|---|---|
| Upper RTI: | J00 **J02·0** **J02·8** J02·9 **J03·0** **J03·8** J03·9 J04·0 J04·1 J04·2 J05·0 J05·1 J06·0 J06·8 J06·9 |
| Lower RTI: | **J12·0** **J12·2** **J12·3** **J12·8** J12·9 **J13** **J14** **J15·0** **J15·1** **J15·2** **J15·3** **J15·4** **J15·5** **J15·6** **J15·7** **J15·8** J15·9 **J16·0** **J16·8** **17·0** **J17·1** **J17·2** **J17·3** **J17·8** J18·0 J18·1 J18·2 J18·8 J18·9 J22 **J20·0** **J20·1** **J20·2** **J20·3** **J20·4** **J20·6** **J20·7** **J20·8** J20·9 |
| Influenza-coded RTI: | **J09** **J10·0** **J10·1** **J10·8** J11·0 J11·1 J11·8 |
| RSV-coded RTI: | **J12·1** **J20·5** **J21·0** |
| COVID-19-coded RTI: | U07·1 U07·2 |
| Respiratory support: | GXAV01 GXAV10 GXAV30 |

Note: Diseases with known pathogen in **bold**. Codes for respiratory support: Ventilation (GXAV01); Continuous Positive Airway Pressure (CPAP) (GXAV10); High flow oxygen therapy (GXAV30).

### **S-Methods:** Estimating projections

Estimation:

First we estimate the number of monthly hospital admissions ($y_t$) using ordinary least square (OLS) regression. The regression model was set up as simple as possible to capture the seasonal variation, i.e. the average hospital admission per calendar month for each calendar year (i.e. $m_1 = 1$ if January if in January otherwise zero, $m_2 = 1$ if in February otherwise zero, ..., $m_{11} = 1$ if in November otherwise zero (December used as reference)), and a possible trend in hospital admissions over time ($k_t$, where $k_1 = 1$ in January 2017, $k_2 = 2$ in February 2017, ..., $k_{36} = 36$ in December 2019 and so on). This is illustrated in the following equation:

$$y_t = \beta_0 + \beta_1 * k_t + \beta_2 * m_1 + \beta_3 * m_2 + ... + \beta_{12} * m_{11} + \epsilon_t$$

From Figure 3 we see that the hospital admissions for the calendar months were very similar in the years 2017 through 2019, and thus this very simple model provides a rough but reasonable representation of the observed seasonal patterns. For three of the four scenarios ("business as usual", "children's immunity debt", and "maternal and child immunity debt") we run the regressions on the observations from January 2017 (t = 1) to December 2019 (t = 36), and used the estimated model — with the amendments described below — to predict the hospital admissions from August 2021 (t = 56) to June 2023 (t = 78) (see Figure 3). For "continuous lockdown" we run the regression on the observations from July 2020 (t = 43) to June 2021 (t = 54), and used the estimated model to predict the hospital admissions from August 2021 (t = 56) to June 2023 (t = 78) (see Figure 3).

Scenario projections:

To get the projected values we forecast the regression for months August 2021 (t = 56) to June 2023 (t = 78) using the following adjustments to the estimated model:

- "Business as usual": No changes.
- "Continuous lockdown": No changes.
- "Children's immunity debt": On top of the predictions from the estimated model, we added the difference between the estimated and observed number of hospital admissions for the period from January 2020 (t = 37) through July 2021 (t = 55), and divided the sum on each calendar month's average share of hospital admissions per season. For example, in January 2021 the predicted number of admissions was 932 and the observed was 132, yielding a difference $d_{Jan21} = 932 - 131 = 800$. The sum over all these differences (d = $d_{Jan20}$ + $d_{Feb20}$ + ... + $d_{Jul21}$) can be considered a simple estimate of the admissions that did not occur ("admission debt") because of the lockdown, and d = 5024. This admission debt was

then distributed to the 2021/22 season by the average share of all admissions in the seasons 2017-2019 that occurred in each calendar month (i.e. the shares that sum 100% were: 17% for January, 16% for February, 12% for March, 7% for April, 6% for May, 5% for June, 2% for July, 3% for August, 6% for September, 7% for October, 8% for November, and 11% for December).

- "Maternal and child immunity debt": We followed the same procedure as in the scenario "Children's immunity debt", except that we did it separately for children aged 0-12 months and 1-5 years. Moreover, the "admission debt" for the children aged 0-12 months ($d_{0-12months} = 2660$) was doubled (i.e. set to $d_{0-12months} = 2 * 2660 = 5320$) before added together with the "admission debt" for children aged 1-5 years ($d_{1-5years} = 2364$) and distributed across the calendar months of the season 2021/2022.

The exact same procedure was used for calculating the projected numbers of hospital admissions with respiratory support. All observed and projected numbers are given in S-Table 2.

### **S-Table 2:** Observed vs. projected RTI-admissions + respiratory support.

| | Hospital admissions | | | | | Respiratory support | | | | |
|---|---|---|---|---|---|---|---|---|---|---|
| Month | Observed | Projected | | | | Observed | Projected | | | |
| | | BAU | CL | CID | MACID | | BAU | CL | CID | MACID |
| Jan-17 | 1144 | | | | | 214 | | | | |
| Feb-17 | 933 | | | | | 158 | | | | |
| Mar-17 | 669 | | | | | 90 | | | | |
| Apr-17 | 427 | | | | | 31 | | | | |
| May-17 | 410 | | | | | 27 | | | | |
| Jun-17 | 346 | | | | | 19 | | | | |
| Jul-17 | 200 | | | | | 17 | | | | |
| Aug-17 | 213 | | | | | 18 | | | | |
| Sep-17 | 402 | | | | | 21 | | | | |
| Oct-17 | 476 | | | | | 28 | | | | |
| Nov-17 | 567 | | | | | 46 | | | | |
| Dec-17 | 600 | | | | | 63 | | | | |
| Jan-18 | 766 | | | | | 95 | | | | |
| Feb-18 | 838 | | | | | 129 | | | | |
| Mar-18 | 807 | | | | | 136 | | | | |
| Apr-18 | 531 | | | | | 78 | | | | |
| May-18 | 398 | | | | | 50 | | | | |
| Jun-18 | 344 | | | | | 31 | | | | |
| Jul-18 | 195 | | | | | 20 | | | | |
| Aug-18 | 275 | | | | | 21 | | | | |
| Sep-18 | 380 | | | | | 31 | | | | |
| Oct-18 | 416 | | | | | 33 | | | | |
| Nov-18 | 488 | | | | | 58 | | | | |
| Dec-18 | 744 | | | | | 111 | | | | |
| Jan-19 | 1027 | | | | | 209 | | | | |
| Feb-19 | 992 | | | | | 211 | | | | |
| Mar-19 | 661 | | | | | 124 | | | | |
| Apr-19 | 385 | | | | | 52 | | | | |
| May-19 | 368 | | | | | 36 | | | | |
| Jun-19 | 280 | | | | | 22 | | | | |
| Jul-19 | 190 | | | | | 14 | | | | |
| Aug-19 | 208 | | | | | 13 | | | | |
| Sep-19 | 374 | | | | | 35 | | | | |
| Oct-19 | 429 | | | | | 34 | | | | |
| Nov-19 | 486 | | | | | 44 | | | | |
| Dec-19 | 609 | | | | | 83 | | | | |
| Jan-20 | 640 | | | | | 89 | | | | |
| Feb-20 | 680 | | | | | 121 | | | | |
| Mar-20 | 451 | | | | | 76 | | | | |
| Apr-20 | 66 | | | | | 2 | | | | |
| May-20 | 77 | | | | | 6 | | | | |
| Jun-20 | 149 | | | | | 13 | | | | |
| Jul-20 | 133 | | | | | 5 | | | | |
| Aug-20 | 193 | | | | | 19 | | | | |
| Sep-20 | 236 | | | | | 14 | | | | |
| Oct-20 | 221 | | | | | 13 | | | | |
| Nov-20 | 218 | | | | | 16 | | | | |
| Dec-20 | 199 | | | | | 8 | | | | |
| Jan-21 | 132 | | | | | 2 | | | | |
| Feb-21 | 151 | | | | | 6 | | | | |
| Mar-21 | 187 | | | | | 9 | | | | |
| Apr-21 | 153 | | | | | 5 | | | | |
| May-21 | 192 | | | | | 10 | | | | |
| Jun-21 | 193 | | | | | 9 | | | | |
| Jul-21 | 159 | | | | | 6 | | | | |
| Aug-21 | 223 | 35 | 258 | 350 | 509 | 12 | 35 | 23 | 78 | 122 |
| Sep-21 | 626 | 47 | 301 | 640 | 883 | 62 | 47 | 18 | 104 | 154 |
| Oct-21 | 1211 | 50 | 286 | 744 | 1045 | 174 | 50 | 17 | 110 | 169 |
| Nov-21 | | 67 | 283 | 883 | 1288 | | 67 | 20 | 149 | 235 |
| Dec-21 | | 104 | 264 | 1143 | 1630 | | 104 | 12 | 229 | 341 |
| Jan-22 | | 197 | 197 | 1763 | 2664 | | 197 | 6 | 419 | 686 |
| Feb-22 | | 190 | 216 | 1651 | 2585 | | 190 | 10 | 405 | 675 |
| Mar-22 | | 141 | 252 | 1249 | 1940 | | 141 | 13 | 300 | 511 |
| Apr-22 | | 78 | 218 | 740 | 1133 | | 78 | 9 | 166 | 277 |
| May-22 | | 62 | 257 | 633 | 914 | | 62 | 14 | 132 | 204 |
| Jun-22 | | 48 | 258 | 501 | 719 | | 48 | 13 | 103 | 162 |
| Jul-22 | | 41 | 264 | 132 | 132 | | 41 | 14 | 41 | 41 |
| Aug-22 | | 42 | 324 | 169 | 169 | | 42 | 28 | 42 | 42 |
| Sep-22 | | 53 | 367 | 322 | 322 | | 53 | 23 | 53 | 53 |
| Oct-22 | | 56 | 352 | 377 | 377 | | 56 | 22 | 56 | 56 |
| Nov-22 | | 74 | 349 | 451 | 451 | | 74 | 25 | 74 | 74 |
| Dec-22 | | 110 | 330 | 588 | 588 | | 110 | 17 | 110 | 110 |
| Jan-23 | | 203 | 263 | 900 | 900 | | 203 | 11 | 203 | 203 |
| Feb-23 | | 196 | 282 | 842 | 842 | | 196 | 15 | 196 | 196 |
| Mar-23 | | 147 | 318 | 634 | 634 | | 147 | 18 | 147 | 147 |
| Apr-23 | | 84 | 284 | 369 | 369 | | 84 | 14 | 84 | 84 |
| May-23 | | 68 | 323 | 313 | 313 | | 68 | 19 | 68 | 68 |
| Jun-23 | | 54 | 324 | 245 | 245 | | 54 | 18 | 54 | 54 |

Note: BAU = Business as usual; CL = Continuous lockdown; CID = Children's immunity debt; MACID = Maternal and child immunity debt.
